# Gene-Temperature Interactions and Risk of Childhood Acute Lymphoblastic Leukemia

**DOI:** 10.64898/2026.07.09.26357608

**Authors:** Tormod Rogne, Rong Wang, Pin Wang, Kai Chen, Shuangge Ma, Joshua L. Warren, Catherine Metayer, Joseph L. Wiemels, Andrew T. DeWan, Xiaomei Ma

## Abstract

**Background:** High ambient temperature in early pregnancy has been linked to an increased risk of childhood acute lymphoblastic leukemia (ALL). To better understand biological mechanisms, the current study evaluated potential interaction between temperature and genetic characteristics.

**Methods:** We used data from California birth records (1982-2008) and California Cancer Registry (1988-2011) to identify ALL cases (n=3,353) diagnosed ≤14 years of age and non-cancer controls (n=3,530) matched 1:1 on sex, race, ethnicity, and birth year and month. Weekly ambient temperatures throughout pregnancy were assessed on a 1-km grid around the birth address, while genetic data were available from a genome-wide association study using neonatal blood spots. We evaluated the association between ambient temperature and ALL risk by quartiles of established genetic risk score for ALL. Next, we formally tested gene-temperature interactions in the association with ALL, correcting for multiple testing, for genes previously identified with epigenetic changes due to both temperature and ALL. All analyses were adjusted for potential confounders.

**Results:** The elevated risk of ALL per 5 °C increase of weekly mean ambient temperature, confined to early pregnancy, was more pronounced among children with the lowest genetic susceptibility to ALL, especially among Latino children (first quartile: odds ratio [OR] = 1.50, 95% confidence interval [CI]: 1.14-1.97); fourth quartile: OR=1.03, 95% CI: 0.83-1.28). There were significant interactions (p<0.002) between ambient temperature and polymorphisms in *BNC1* among non-Latino White children, and suggestive interactions (p<0.05) with *TBPL2* and *NRXN1* in the full population.

**Conclusions:** Our findings suggest that there may be interactions between ambient temperature in early pregnancy and offspring genotype in the risk of childhood ALL.

**Impact:** If replicated, these findings could help elucidate the biological mechanisms linking high ambient temperature in early pregnancy and the risk of childhood ALL.

## INTRODUCTION

Increasing ambient temperatures due to climate change have been linked to a variety of adverse health outcomes.^1^ One such potential adverse outcome is childhood acute lymphoblastic leukemia (ALL), which is the most common childhood malignancy.^2^ We previously observed a significant association between high ambient temperature during pregnancy and an increased risk of ALL in children.^3^ Possible mechanisms for this association, however, are unclear. It is probable that genetic variations may affect the vulnerability to causative environmental factors, exhibited as gene-environment interactions.^4^ Such interaction analyses may also identify genetic loci that do not have marginal effects on the outcome. While several studies have evaluated the links between air pollution and risk of cancer by levels of genetic susceptibility to that cancer, there have been no such studies for ambient temperature.^5,6^

The aims of this study were twofold. Using a genotyped subsample of our previous study on temperature and ALL risk,^3^ we first evaluated whether ambient temperature was differentially associated with risk of ALL depending on the subjects’ genetic susceptibility to ALL. Next, we tested specific gene-temperature interactions using a set of genes previously reported to exhibit epigenetic changes due to both ambient temperature and ALL (indicative of genetic regions relevant to this exposure-outcome association).

## METHODS

### Study Population

This study used data from the California Childhood Cancer Record Linkage Project (CCRLP), which linked statewide birth records from the California Department of Public Health with cancer diagnoses reported to the California Cancer Registry. The current project used data for the years of birth when genetic data was available from 1982 to 2008, and cancer diagnosis were available from the registry between 1982 and 2011.^7^ This study received approval from the institutional review boards at the California Department of Public Health, Yale University, University of California, Berkeley, and University of Southern California.

Cases were defined as children diagnosed with ALL at 0-14 years of age, including the International Classification of Diseases for Oncology, 3^rd^ edition, codes 9820, 9826, 9835-9837 and 9948. Subjects not identified with any cancer diagnosis were eligible as controls, and they were randomly selected from the birth records and 1:1 matched to cases on sex, race, ethnicity, and birth year and month. Of 7,203 genotyped and geocoded subjects, we excluded 416 subjects with missing data on gestational duration, 12 subjects with missing temperature data, 13 subjects with missing data on neighborhood poverty index, and one subject with missing birthweight data; totaling a final study sample of 6,883 subjects, of which 3,353 were cases and 3,530 were controls.

### Genetic Data

Genotyping was conducted with the Affymetrix Axiom World Array using DNA extracted from neonatal blood spots. Ancestry-informed principal components were generated by Eigenstrat. Further details and quality control has previously been described.^7^ For the current project, the directly genotyped data were further imputed using the TOPMed Imputation Server.^8^ The TOPMed r2 reference panel was used, with Eagle v2.4 phasing.

The genetic risk score (GRS) for childhood ALL was constructed based on a genome-wide association study (GWAS) of 5,321 childhood ALL cases and 16,666 controls of European ancestry, the largest GWAS on childhood ALL to date not overlapping with any subjects in CCRLP.^9^ All of the 15 independent (r^2^<0.01) single-nucleotide polymorphisms (SNPs) strongly (p-value<5e-8) associated with ALL that were identified in the GWAS were used to construct the GRS. The log odds ratio (OR) of the association between the effect allele and risk of ALL in the original GWAS was used as a weight in the GRS estimation.

### Temperature Ascertainment

For all cases and controls, maternal residential addresses at the time of birth were geocoded.^10^ Daily minimum and maximum temperature data were collected from Daily Surface Weather Data on a 1-km Grid for North America, Version 4.^11^ Daily means were estimated from the average of daily minimum and maximum temperatures. The exposure of interest in this study was mean weekly temperature in each gestational week, calculated as the 7-day mean of the daily means. Associations are reported based on a 5 °C increase in mean weekly ambient temperature, roughly corresponding to a one standard deviation (SD) increase, as in our previous analysis.^3^

### Other Variables of Interest

Data on parity, maternal age and level of education, offspring sex, birthweight and gestational age at delivery, were obtained from the birth records. Date of the first day of last menstrual period was estimated based on date of birth and registered gestational age at birth. By linking maternal residential addresses to the 2000 census, we obtained data on the percentage of people living below the 150% poverty level at the block group level. Data on Californian counties and regions were collected from the California Department of Health Care Services, Medi-Cal County Code Reference Table.^12^ Ancestry was inferred from the genotyped data and divided into Latino, non-Latino White, non-Latino Black, non-Latino Asian and Other.^7^

### Statistical Analyses

We evaluated the association between weekly mean ambient temperature and risk of childhood ALL in each gestational week separately. We considered the weeks from 12 weeks prior to the first day of the last menstrual period through gestational week 22 to capture the exposure window previously identified.^3^ ORs and 95% confidence intervals (CIs) were estimated in multivariable unconditional logistic regression models that adjusted for the matching variables and other covariates.^13^ Specifically, we adjusted for sex, birth year, season of conception (winter [December, January, and February], spring [March, April, and May], summer [June, July, and August], and fall [September, October, and November]), maternal age (≤19, 20-24, 25-29, 30-34, ≥35 years), birth order (first, second, third or higher), maternal education (8^th^ grade or less, 9-12^th^ grade, at least some college, unknown), neighborhood poverty (in tertiles), county of residence (58 counties), and principal components 1 to 10. The analyses were first conducted including all subjects in the same model, adjusting for ancestry. Additionally, analyses were carried out separately for Latino and non-Latino White children, while there were insufficient number of subjects in the other ancestry groups for subgroup analyses. After identifying the gestational week with the most pronounced association between ambient temperature and risk of ALL, all subsequent analyses were based on that week. In the interaction analyses by genetic susceptibility to ALL, the GRS was grouped into quartiles, and analyses were run within strata of the GRS quartile. Differences between the subgroups were formally tested by the Z-statistic.^14^

In order to conduct a hypothesis-free test of interactions between specific genes and temperature in relation to the risk of ALL, we first identified a set of SNPs to test, as we would not have the power to test all SNPs in the dataset. We used the “meet-in-the-middle” approach where we investigated the overlap between exposure-associated (i.e., temperature) and outcome-associated (i.e., childhood ALL) methylation changes.^15^ For ambient temperature, we identified three studies with epigenome-wide evaluations of methylation changes due to ambient temperature,^16–18^ while we used data from two epigenome-wide association studies of childhood ALL to identify relevant methylation changes for that outcome.^19,20^ After identifying genes with methylation changes due to both temperature and ALL, the next step was to identify a single tag-SNP for each gene. To achieve this, we used full summary statistics from a previous GWAS on childhood ALL that used the California Childhood Leukemia Study;^7^ this does not overlap with the CCRLP dataset, thus reducing the risk of inflated effect estimates.^21^ For each identified gene, we selected the SNP that was the most strongly associated with risk of ALL based on the p-value of the association. The interaction analyses were then run adjusting for the same set of covariates as in the other analyses.

The p-value threshold for statistical significance was set at 0.05 for most analyses. In the hypothesis-free gene-specific analyses, however, we applied a more stringent Bonferroni-corrected threshold for statistical significance of 0.05 divided by the number of genes tested; p-values below 0.05 but above the Bonferroni-corrected threshold were considered suggestive.

### Software

Plink version 1.9 was used to calculate the GRS and to run the temperature-SNP interaction analyses.^22^ For the remaining analyses, R version 4.2.0 was used, applying the packages rgdal for attaching county and region to the geographic coordinates and biomaRt to annotate the SNPs and genes.

## RESULTS

Among the 6,883 children in the study, the majority were Latino American and male (Table 1). As per matching design, sex, race, ethnicity, year of birth, and season of conception were equally distributed between cases and controls. Of the seven Californian regions, both cases and controls were most likely to live in the Los Angeles region. Cases, compared with controls, were more likely to have older mothers and to have a heavier birth weight.

**Table 1.**
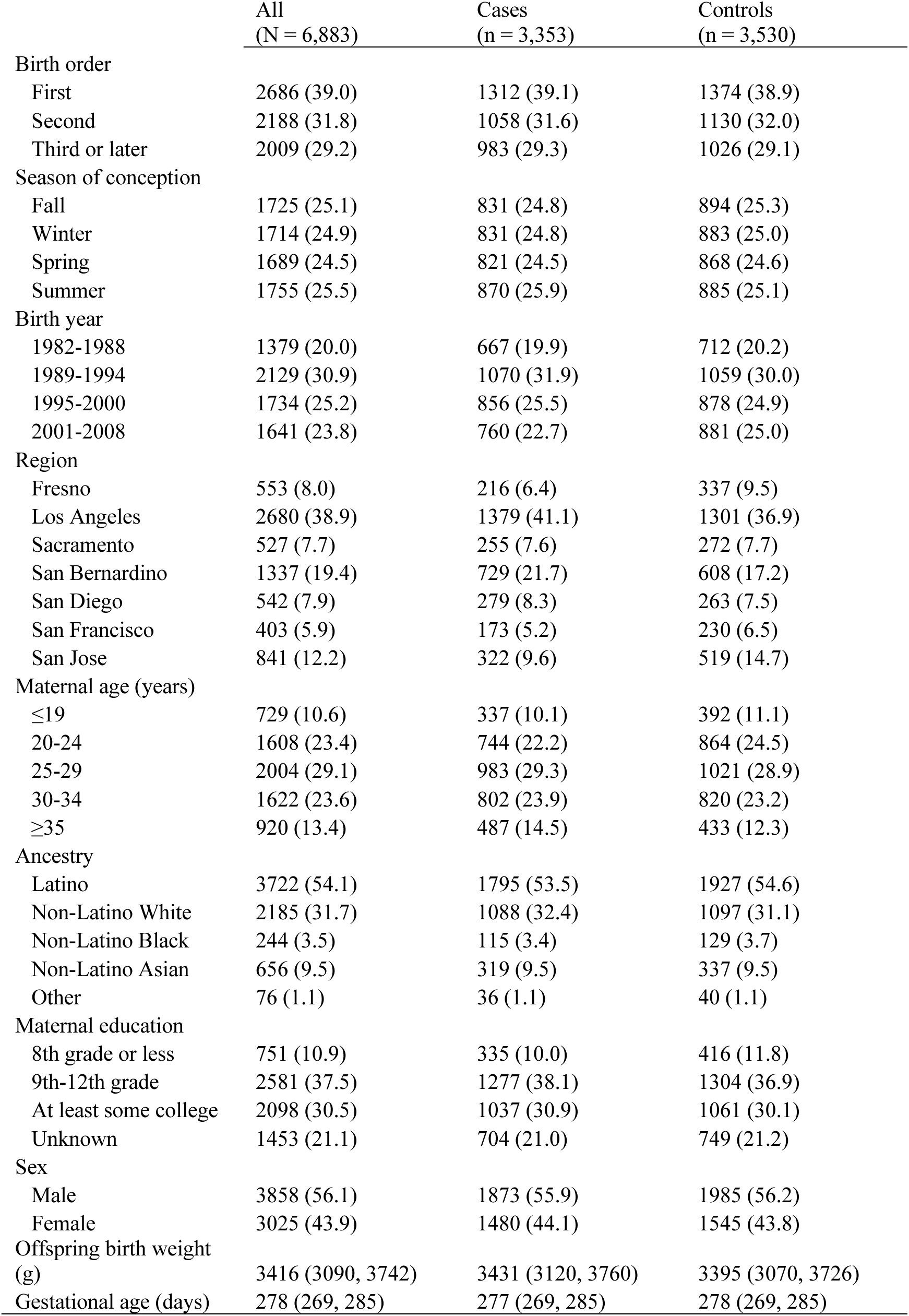
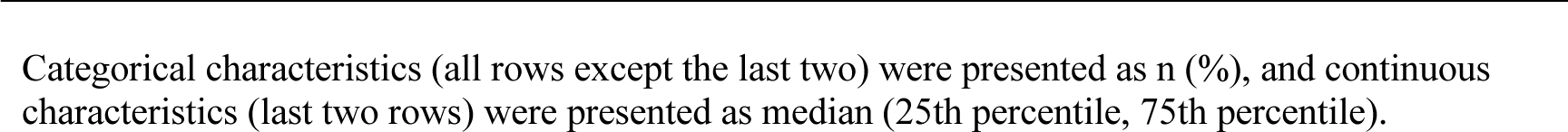
Characteristics of the study population.

|  | All<br>(N = 6,883) | Cases<br>(n = 3,353) | Controls<br>(n = 3,530) |
| --- | --- | --- | --- |
| Birth order |  |  |  |
| First | 2686 (39.0) | 1312 (39.1) | 1374 (38.9) |
| Second | 2188 (31.8) | 1058 (31.6) | 1130 (32.0) |
| Third or later | 2009 (29.2) | 983 (29.3) | 1026 (29.1) |
| Season of conception |  |  |  |
| Fall | 1725 (25.1) | 831 (24.8) | 894 (25.3) |
| Winter | 1714 (24.9) | 831 (24.8) | 883 (25.0) |
| Spring | 1689 (24.5) | 821 (24.5) | 868 (24.6) |
| Summer | 1755 (25.5) | 870 (25.9) | 885 (25.1) |
| Birth year |  |  |  |
| 1982-1988 | 1379 (20.0) | 667 (19.9) | 712 (20.2) |
| 1989-1994 | 2129 (30.9) | 1070 (31.9) | 1059 (30.0) |
| 1995-2000 | 1734 (25.2) | 856 (25.5) | 878 (24.9) |
| 2001-2008 | 1641 (23.8) | 760 (22.7) | 881 (25.0) |
| Region |  |  |  |
| Fresno | 553 (8.0) | 216 (6.4) | 337 (9.5) |
| Los Angeles | 2680 (38.9) | 1379 (41.1) | 1301 (36.9) |
| Sacramento | 527 (7.7) | 255 (7.6) | 272 (7.7) |
| San Bernardino | 1337 (19.4) | 729 (21.7) | 608 (17.2) |
| San Diego | 542 (7.9) | 279 (8.3) | 263 (7.5) |
| San Francisco | 403 (5.9) | 173 (5.2) | 230 (6.5) |
| San Jose | 841 (12.2) | 322 (9.6) | 519 (14.7) |
| Maternal age (years) |  |  |  |
| ≤19 | 729 (10.6) | 337 (10.1) | 392 (11.1) |
| 20-24 | 1608 (23.4) | 744 (22.2) | 864 (24.5) |
| 25-29 | 2004 (29.1) | 983 (29.3) | 1021 (28.9) |
| 30-34 | 1622 (23.6) | 802 (23.9) | 820 (23.2) |
| ≥35 | 920 (13.4) | 487 (14.5) | 433 (12.3) |
| Ancestry |  |  |  |
| Latino | 3722 (54.1) | 1795 (53.5) | 1927 (54.6) |
| Non-Latino White | 2185 (31.7) | 1088 (32.4) | 1097 (31.1) |
| Non-Latino Black | 244 (3.5) | 115 (3.4) | 129 (3.7) |
| Non-Latino Asian | 656 (9.5) | 319 (9.5) | 337 (9.5) |
| Other | 76 (1.1) | 36 (1.1) | 40 (1.1) |
| Maternal education |  |  |  |
| 8th grade or less | 751 (10.9) | 335 (10.0) | 416 (11.8) |
| 9th-12th grade | 2581 (37.5) | 1277 (38.1) | 1304 (36.9) |
| At least some college | 2098 (30.5) | 1037 (30.9) | 1061 (30.1) |
| Unknown | 1453 (21.1) | 704 (21.0) | 749 (21.2) |
| Sex |  |  |  |
| Male | 3858 (56.1) | 1873 (55.9) | 1985 (56.2) |
| Female | 3025 (43.9) | 1480 (44.1) | 1545 (43.8) |
| Offspring birth weight<br>(g) | 3416 (3090, 3742) | 3431 (3120, 3760) | 3395 (3070, 3726) |
| Gestational age (days) | 278 (269, 285) | 277 (269, 285) | 278 (269, 285) |

Categorical characteristics (all rows except the last two) were presented as n (%), and continuous characteristics (last two rows) were presented as median (25th percentile, 75th percentile).

The GRS for ALL explained 7.2% of the variation of ALL risk in the full sample. Assessing each genetic ancestry group separately, the explained variance was 7.3%, 9.1%, 1.8%, 4.8% and 9.7% for Latino, non-Latino White, non-Latino Black, non-Latino Asian and Other, respectively.

In the adjusted model, the most pronounced association between ambient temperature and risk of ALL was observed in gestational week 0, where a 5 °C increase was associated with an OR for ALL of 1.12 (95% CI 1.04 to 1.21) (Supplementary Figure 1). The mean of the weekly mean ambient temperature in gestational week 0 was 17.5 °C (SD 5.1 °C) for the full study population, 17.8 °C (SD 5.0 °C) among cases and 17.3 °C (5.1 °C) among controls.

In stratified analyses by the GRS, we observed that in gestational week 0, high ambient temperature was most strongly associated with odds of ALL among those with low genetic risk for ALL (Figure 1), but the difference between the first (OR 1.31, 95% CI 1.11 to 1.54) and fourth (OR 1.11, 95% CI 0.94 to 1.31) quartiles was not statistically significant (p-value = 0.171). The pattern was more pronounced among Latino children, where the difference between the first (OR 1.43, 95% CI 1.09 to 1.87) and fourth (OR 1.03, 95% CI 0.83 to 1.28) quartiles was close to statistically significant (p-value = 0.061).

**Figure 1.**
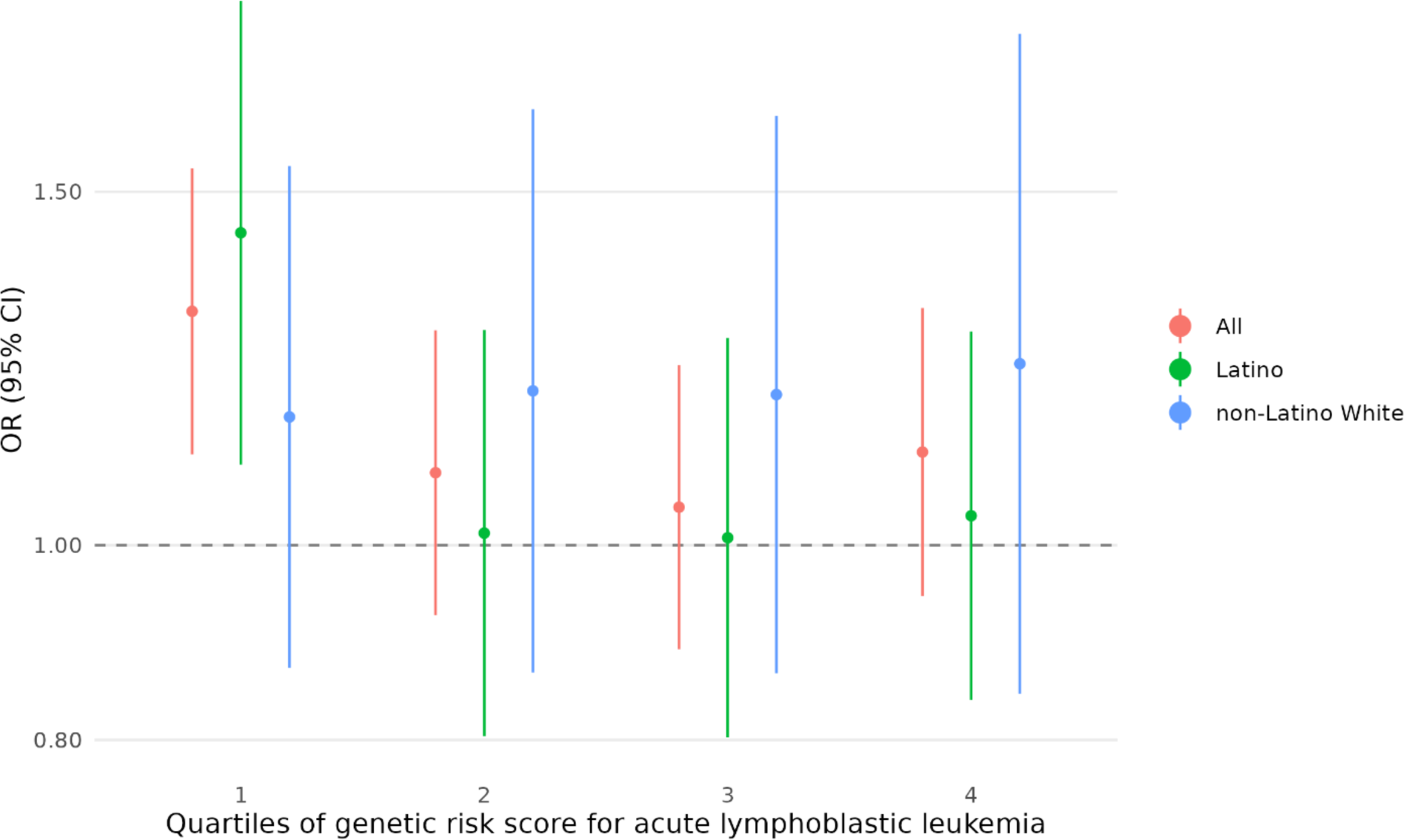
Ambient temperature in gestational week 0 and risk of childhood acute lymphoblastic leukemia by genetic risk score for acute lymphoblastic leukemia. OR per 5 °C increase in mean weekly ambient temperature in gestational week 0 by quartiles of the GRS for ALL. Lowest quartile corresponds to the lowest genetic risk of ALL. Adjusted for sex, birth year, season of conception, maternal age, birth order, maternal education, neighborhood poverty, county of residence, and principal components 1 to 10. CI: confidence interval; OR: odds ratio.

Using the meet-in-the-middle approach, we identified 2,172 genes associated with epigenetic changes due to childhood ALL and 131 genes associated with epigenetic changes due to ambient temperature. Of these genes, 21 were affected by both ALL and temperature. In the analysis of all ancestries combined, there were no statistically significant interactions (p-value threshold 0.05/21 = 0.002) between ambient temperature in gestational week 0 and the marker SNP of any of the 21 genes (Table 2), although two genes (*TBPL2* and *NRXN1*) were suggestive. In the non-Latino White only analysis, there was a statistically significant interactive association with *BNC1*. There were no significant or suggestive associations in the Latino only analysis.

**Table 2.**
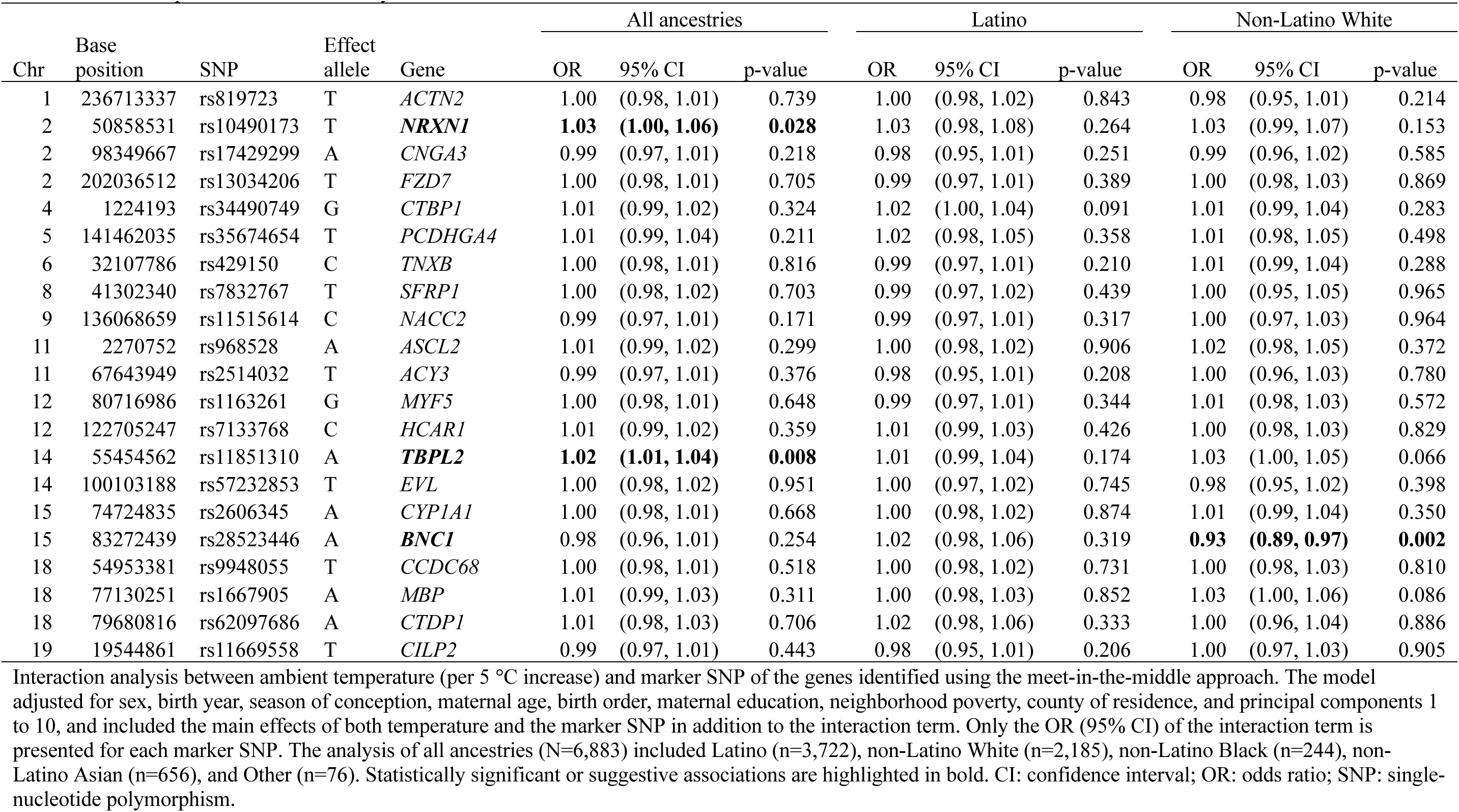
Gene-temperature interaction analyses.

## DISCUSSION

In the first study to evaluate gene-temperature interactions in the etiology of childhood ALL, we observed that the association between high ambient temperature and risk of ALL tended to be more pronounced among subjects at the lowest genetic risk of ALL. Furthermore, we identified a significant interaction between temperature and *BNC1* among non-Latino White children, and suggestive interactions with *NRXN1* and *TBPL2* in the full study population.

While there have been no previous gene-temperature interaction studies on risk of cancer, there have been a few interaction studies on air pollution.^5,6^ These studies have generally observed the association between the exposure and the outcome to be more pronounced among those with the highest genetic susceptibility to the outcome. Contrary to this, we observed that those at the lowest genetic risk of ALL had the most pronounced association between ambient temperature and risk of ALL, albeit not statistically different from the other genetic risk groups. While we do not have a clear explanation for this observation, we hypothesize that non-genetic pathways to ALL may play a relatively larger role among those at low genetic risk of the disease. This was particularly true for Latinos, which – following the same hypothesis – may be due to Latino pregnant women in California carrying a greater risk of personal exposure to high ambient temperature compared with non-Latino White women e.g., due to occupation or residential segregation.^23,24^

At the single-gene level, there was a significant interaction between ambient temperature and *BNC1* in the non-Latino White population. The gene, which encodes for a zinc finger protein, is often inactivated among children with ALL.^25^ Additionally, it is involved in the regulation of keratinocyte proliferation in epidermis and hair follicles, which in turn are functional in thermoregulation.^26,27^ In the full study population, there were suggestive interactions between ambient temperature and the genes *NRXN1* and *TBPL2* in the risk of ALL. *NRXN1* encodes for the cell surface protein Neurexin 1, which is involved in nervous system function and cell-cell-interactions in synapses and endocrine cells.^28^ TBPL2, encoded from *TBPL2*, is located in the cytoplasm and nucleus, and is involved in multiple biological pathways.^29^ One pathway that seems particularly relevant in this context is that of gastrulation. This is a key morphogenic remodeling event in embryonic development that starts around 14 days after fertilization and ends a week later,^29,30^ which aligns well with the susceptible window identified in the current study as well as our previous study^3^.

This study has several strengths but also some limitations. We used state-wide, population-based birth records and cancer registry data to identify cases and controls and obtain information related to pregnancy and birth, minimizing selection bias, outcome misclassification and recall bias. Accurate temperature data on a small geographic aggregation (i.e., 1-km grid) was used. An important limitation is that the temperature exposure was based on maternal residential address at the time of birth and did not account for access to air conditioning, travel, or type of work; thus, an individual’s true exposure to ambient temperature was unknown. Any exposure misclassification, however, is likely to be non-differential and biasing towards the null. Also, cases and controls were matched on birth month and year, which could attenuate associations between ambient temperature and risk of ALL. Time-invariant confounding is unlikely given that we observed a specific critical exposure window,^31^ which also aligned with our previous findings.^3^ We did not have the statistical power to pursue gene-temperature interaction analyses genome-wide, and our analysis comparing the temperature-ALL associations by quartiles of GRS for ALL was slightly underpowered; still, our sample size was substantial for a gene-environment interaction study of a rare childhood outcome. The GRS was constructed based on findings of a large GWAS that predominantly included subjects of European ancestry, so it may not be the most informative for individuals with other ancestries. Still, the explained percentage of variance in Latino children (7.3%) was not much worse than that in non-Latino White Children (9.1%).

In conclusion, we observed interactive effects between offspring genetics and prenatal ambient temperature exposure in the risk of childhood ALL. Follow-up studies in independent populations are encouraged so that we can better understand the potential biological links between high ambient temperature and ALL risk in children.

## Data availability

The authors are unable to share data presented in this manuscript, as we are prohibited by California statutes from publicly sharing information that is derived from data and biospecimens of the California Department of Public Health. We welcome questions from other investigators or requests for additional analyses that are pertinent to the data presented in this manuscript. Other investigators can apply for the same data from the Committee for the Protection of Human Subjects at the California Health and Human Services Agency, as we did for this study.

## Acknowledgements

The collection of cancer incidence data used in this study was supported by the California Department of Public Health pursuant to California Health and Safety Code Section 103885; Centers for Disease Control and Prevention’s (CDC) National Program of Cancer Registries, under cooperative agreement NU58DP007156; the National Cancer Institute’s Surveillance, Epidemiology and End Results Program under contract HHSN261201800032I awarded to the University of California, San Francisco, contract HHSN261201800015I awarded to the University of Southern California, and contract HHSN261201800009I awarded to the Public Health Institute. The CCRLP GWAS was primarily funded by the National Cancer Institute (R01CA155461) with additional support from the National Institute of Environmental Health Sciences (P50ES018172) and the United States Environmental Protection Agency (EPA; assistance agreement RD83615901). For the GWAS summary statistics from the California Childhood Leukemia Study, we utilized data from the California Biobank Program (SIS request #26). The ideas and opinions expressed herein are those of the authors and do not necessarily reflect the opinions of the State of California, Department of Public Health, the National Cancer Institute, the EPA, and the Centers for Disease Control and Prevention or their Contractors and Subcontractors.

