## Supplementary Figure 1 for "Gene-Temperature Interactions and Risk of Childhood Acute Lymphoblastic Leukemia"

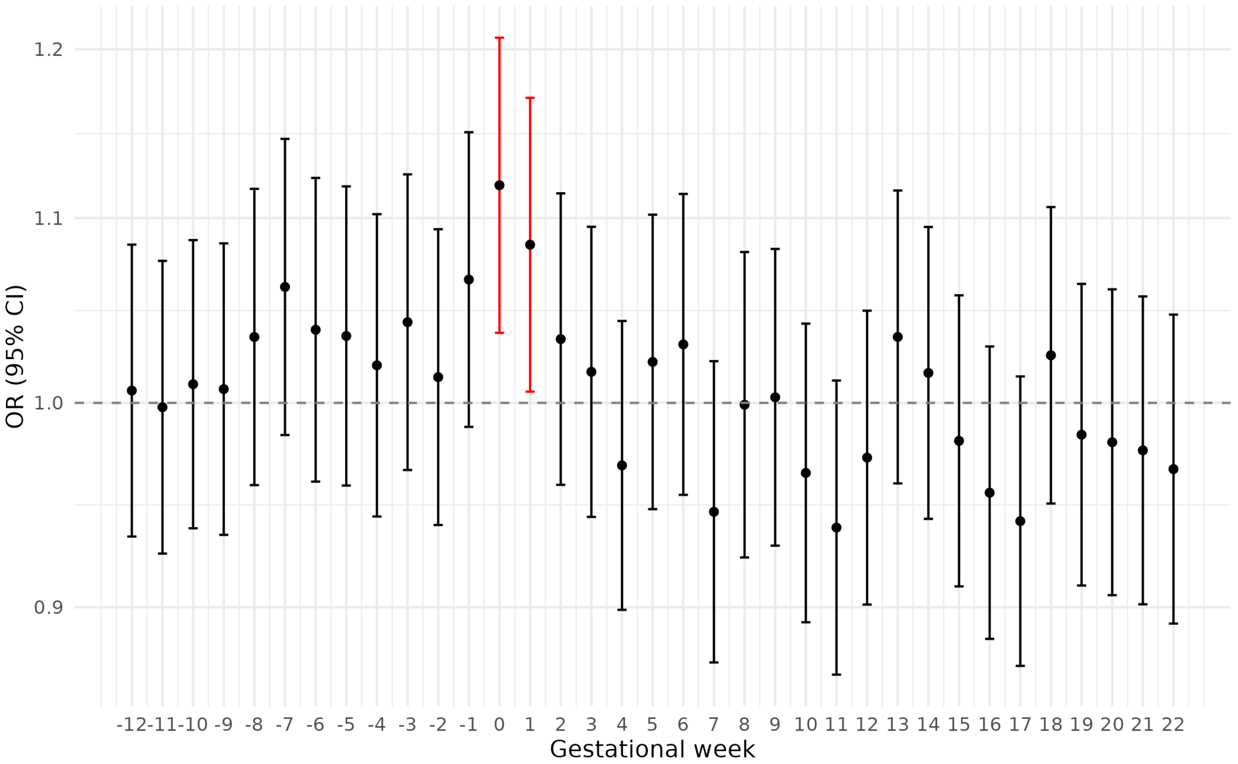


**Supplementary Figure 1.** Gestational week-specific associations between ambient temperature and risk of childhood acute lymphoblastic leukemia.

*Legend:* Results from gestational week-specific analyses of the association between ambient temperature and risk of childhood acute lymphoblastic leukemia. Each gestational week was analyzed separately, adjusting for sex, birth year, season of conception, maternal age, birth order, maternal education, neighborhood poverty, county of residence, and principal components 1 to 10. Unit of exposure per 5 °C increase in mean weekly ambient temperature. Statistically significant associations are shown in red. CI, confidence interval; OR, odds ratio.
